# Social and labor stressors influencing disability in Adult Attention Deficit/Hyperactivity

**DOI:** 10.1101/2021.12.12.21265631

**Authors:** Dimitri Marques Abramov, Marjorie Mastellaro Baruzzi, Renata Joviano Alvim, Ana Carolina Moda Nunes Peixoto, Victor de Souza Mannarino, Caroline Barros Pacheco Loureiro, Danilla Ferreira, Iara Almeida, Ingrid Pinheiro, Rosângela Marques Valentim

## Abstract

Attention Deficit/Hyperactivity Disorder (ADHD) is a controversial issue. If ADH represents a mental disorder, it must be the cause of a primary dysfunctionality and maladaptation from childhood to adult life. We will look for evidence to substantiate this discussion. We conducted an online survey about economic and academic performances and maladaptation, following a screening for ADHD using Adult Self Report Scale (ASRS). The subjects were naive. There were 2173 participants, of which 28.06% were ADH(+). Even regarding only subjects with extreme ASRS scores (<1.0 and >2.5), ADH(+) and (−) groups did not shown difference in functionality. We grouped subjects by professional career. The highest ADH(+) prevalence was found in publicity, where almost no difference in subjective suffering between the groups was observed. Our results indicate that ADH(+) people can show equivalent functionality and adaptability than ADH(−) ones when they live in their preferred labor/social niches, arguing that ADHD can be a different cognitive style with dysfunctionality and mental suffering could be secondary to social stress.

## Introduction

An appropriate definition of mental disorder is a maladaptive process secondary to the failure of an underlying system to perform functions as determined by Evolution [1]. Under this rationale, the DSM-5 defines mental disorder as the phenomenological manifestation of a dysfunctional and maladaptive mental process [2]. Exhaustive epistemological discussions about the determinants of this nosology have already lasted decades, since it is argued that the normative, cultural, and moral constructs of our world define what mental disorders are [1,3]. However, individual diversity would manifest incompatibilities and consequent suffering with a system of finite and hegemonic norms [4]. The relationship between mental suffering in people with a psychiatric diagnostic and social stress is known. Several consequences are linked to mental health-related stigma and discrimination in patients, e.g. loss of self-esteem, self-efficacy, and negative responses such as fear, rage, or guilt [5]. Interventions have been proven to reduce mental illness related to stigma and discrimination [6, 7].

A major source of discussion throughout society is Attention Deficit Hyperactivity Disorder (ADHD). Despite the evidence that ADHD is a well-defined mental phenomenology by biological bases [8–10], its pathological character is quite controversial, both inside academic walls [11,12] and according to common sense [13]. Nevertheless, ADHD is organized as a mental disorder in DSM while there is significative impairment [2]. Therefore, this impairment might be related to the inability of the social system to hold cognitive diversity.

Far beyond these discussions, it is well known that people who manifest ADHD in childhood or adulthood suffer lifelong consequences. Several studies correlate ADH with risky and impulsive behaviors as well as sociopathic profile [14, 15], higher incidence of drug addiction [16], and mostly some impairment in social, economic, and academic functioning [17, 18]. However, a more complex approach would be required to discuss if these findings are primarily inherent to ADHD or if they could be secondary to the social stress to which people have been exposed since childhood.

Through naive participants screened as ADH(+) or ADH(−) by ASRS, our goal was to verify whether academic and economic performance as well as the manifestation of subjective suffering (related to work or not) were different in these two populations. We also accessed these dimensions by professional careers to verify if in some context ADH(+) people show functionality and adaptability equivalent to ADH(−) ones.For instance, if there are working/professional environments where ADH(+) subjects show functionality and adaptability equivalent to typical people, we have an indirect evidence that ADHD, as described by DSM-5 [2], could not be a proper mental disorder but alternatively a circumstantial condition of mental suffering, secondary to maladaptive social and work stress since childhood.

## Methods

### Design

We conducted an online survey of an on-demand sampling (people in the authors’ social networks were indistinctly invited to participate via Facebook and WhatsApp). The participants were not aware about ADH evaluation (they are naïve). They answered a self-assessment questionnaire containing the six items from the short ASRS v1.1. version, the WHO assessment for ADHD screening [19–22]. Subjects also answered questions about scholarship, financial incomes, work satisfaction, self-perception of clinical and mental problems and psychic treatments. Data were analyzed regarding several professional niches, delimited by different careers. Due to the methodological limitations inherent to this study and its exploratory nature, our conclusions were drawn as preliminary findings.

### Subjects

A total of 2173 participants (older than 18 y.o.), fluent in Portuguese, were recruited on demand through social networks (Facebook and WhatsApp). They were invited to answer a questionnaire on “Relationships between Behavior, Health, and Working Life” available on Google Forms website (docs.google.com/forms). The electronic informed consent is the first screen of the questionnaire.

The invitation to participate did not mention ADHD. Therefore, all participants were naive. Participation was completely anonymous and there was no link with the participants’ identities. This work and the procedures adopted were duly approved by the Research Ethics Committee of Fernandes Figueira National Institute of the Woman, Child and Adolescent Health (CEP-IFF/FIOCRUZ), registered by CAAE: 80765917.7.0000.5269, under national laws and in accordance with the ethical standards laid down in the 1964 Declaration of Helsinki and its later amendments.

### Questionnaire and its application

The questionnaire had 45 questions in Portuguese, informing about age, sex, self-perceived gender, objective aspects of their lives (demographical information) as well their subjective self-perception about work and health. These answers are our major interest in the survey. “functionality” was estimated according to academic and economic performances (highest level of education and current monthly incomes). We also asked about suffering related to work, and in life in general, through self-perception of clinical and psychic problems, and history of psychotherapeutic and related interventions. We showed a list with 96 professional careers to be freely marked.

ASRS is shown in questions 15 to 20, which are formatted according to the Likert scale (never / seldom / sometimes / often / very often – Likert scale). Following ASRS, many distractor questions were presented to mask the re-presentation of ASRS questions number 1, 2, 4, 5, and 6 into questions 29, 37, 32, 35, and 45, respectively (to infer directly about the individual consistency of answers). Time to complete the questionnaire was not counted. The form did not induce forced responses, i.e., the subjects could choose not to answer any questions. Table 1 shows a summary of the questionnaire used.

**Table 1.** Self-appraising questionnaire.

| no | Question <sup>(I)</sup> | Answer |
| --- | --- | --- |
| 1 | how old are you (in years)? | Entering two digits |
| 2 | What is your biological sex (as your body born) | Male/Female |
| 3 | What is your gender? | Man/Woman/Not binary |
| 4 | Which is highest schooling level that you have? | Elementary, high school (incomplete), high school (complete), graduation (incomplete), graduation (complete), post graduation, none |
| 5 | Which college (s) have you completed or are you still studying (if you have one)?* | *See suppl informaiton 01* |
| 6 | Have you completed or are taking a technical course? If so, check the option (s) that best fits your course (s)* | *See suppl information 01* |
| 7 | What the area of your profession? | Profession of my education (college, technical course, postgraduate); Professions using communication and relationship: Teacher, politician, actor, sales promoter, etc<br>Professions using manual skills and creativity: craftsman, hairdresser, tattoo artist, cheff, designer, fine arts, advertising creation etc<br>Administrative, legal and accounting professions: banking, securities broker, secretary, accountant, notary, forwarder, etc<br>Security Professions: Police, Private Security, City Guard, Military<br>Professions with specific skills: equipment maintenance, mechanic, bricklayer, turner, electrician, driver, masseur, etc<br>Other profession (from another area and / or does not require specific skill)<br>I have no profession |
| 8 | How much are your montly incomes (in reals)? | 11 levels varying in R\$ 1500,00 untill “R\$ 15.000,00 or more” |
| 9 | Why did you choose your profession?* | Vocation, money/market /prestige, family desire, opportunity, lack of choice, I thought it would be easy |
| 10 | Would you change your profession? | Yes/No |
| 11 | Have/Had you some health problem? Which?* | Heart disease, respiratory, bones and joints, neurological, diabetes, high blood pressure, strong allergies, others, none |
| 12 | Have/had you some psychic problem (mental health)? which?* | Anxiety, panic attack, phobias, OCD, depression (1 ep), recurrent depression, chronic depression, BD, schizophrenia, ADHD, asperger, other, none. |
| 13 | Have you ever done or do psychologist (or similar) therapy? | Yes/No |
| 14 | Please indicate which of the following best reflects how you feel about your work: | Usually brings me pleasure and happiness / Usually causes me psychological suffering. |
| 15 | How often do you have trouble wrapping up the fine details of a project, once the challenging parts have been done? | Never<br>seldom<br>Sometimes<br>often<br>Very often |
| 16 | How often do you have difficulty getting things in order when you have to do a task that requires organization? |  |
| 17 | When you have a task that requires a lot of thought, how often do you avoid or delay getting started ? |  |
| 18 | How often do you have problems remembering appointments or obligations ? |  |
| 19 | How often do you fidget or squirm with your hands or your feet when you have to sit down for a long time? |  |
| 20 | How often do you feel overly active and compelled to do things, like you were driven by a motor? |  |
| 21 | How often do you feel insecure about commitments or tasks that you know you are able to solve or are prepared to accomplish? |  |
| 22 | How often do you feel unwell in crowded places? |  |
| 23 | How often do you find it difficult to stand in line and wait for your time? |  |
| 24 | How often do you have an excessive need for sexual stimulation (from pornography to sexual intercourse)? |  |
| 25 | How often do you worry excessively about problems that aren't that big? |  |
| 26 | When you start drinking, how often do you have difficulty stopping? (if you don't drink, mark "never") |  |
| 27 | Do you find it easy to make new friends or meet new people? |  |
| 28 | How often do you find a creative solution to everyday problems, making them easier to solve? |  |
| 30 | Do you currently use psychoactive substances other than alcohol (marijuana, cocaine, stimulants)? |  |
| 31 | In the PAST, did you use psychoactive substances other than alcohol (marijuana, cocaine, stimulants)? In times of heavy consumption, how often did you use these substances? |  |
| 33 | How often do you find it difficult to control the use of any substance that has an effect on your mind (alcohol, tobacco, drugs, tranquilizers)? |  |
| 34 | How often do you feel a deep emptiness, as if life makes no sense? |  |
| 36 | How often do you easily get emotional (sadness, fear, anger, etc.) in situations where other people usually do not respond so strongly? |  |
| 38 | How often do you have a good night's sleep without having to use medications, drugs or the like? |  |
| 39 | How often do you feel excessively needy, in need of being loved? |  |
| 40 | How often during your childhood did adults in your family fight with each other, with other people, or with you, using physical or verbal |  |
|  | violence? |  |
| 41 | How often in childhood and adolescence did you suffer social rejection or were you slighted because of your way of being? |  |
| 42 | How often do you have difficulty sleeping naturally (without using medications, teas or drugs)? |  |
| 44 | How often were your parents understanding and supportive of your difficulties and your limits (including school)? |  |
| 45 | How often do you use any chemicals (alcohol, tobacco, drugs or tranquilizers) when you feel nervous, anxious, distressed, etc.? |  |

(\*): questions that allow more than one answer simultaneously.
Many answers have been simplified because of table sizing. Original questionnaire (in Portuguese), please contact the authors.

ASRS v1.1 short version is highlighted. The redundant or repeated questions are suppressed. Questions from 38 and 42 are opposing, for index I.

### Data analysis

The main analyses were based on screening according to ASRS (short version with six questions) [19, 22], defined subjects as ADH(+) or ADH(−). However, we also made inferences on differences between males and females aiming a description framework between two kinds of people whose differences have no pathological meaning.

According to the ASRS score, to classify as ADH(+), the score must be equal to or higher than 1.6. To calculate it, questions 1 to 3 have a weight of 1.2, while questions 4 to 6 have a weight of 0.8 [22]. The ASRS score adopted here is the average of the weighted scores of the six questions.

The scaled answers (questions 4 and 8) were quantified by subject; from 0 to 7 for schooling level and from 0 to 11 for incomes (see table 1). In question 4, we regarded “technical education” (level 5) as higher to high school because this modality encompasses high school in Brazil.

In a preliminary analysis, we observed a significant correlation between age and incomes (see results). Therefore, to normalize income by eliminating age bias, we created a derived variable that is *income*. The number of clinical and psychic problems per participant were computed as three new quantitative variables.

To infer reliability of ASRS answers, which form a small and two-dimensional cluster (4 questions for inattention and 2 for hyperactivity / impulsivity), Cronbach’s alpha index was calculated [23,24]. We regarded α ≥ 0.60 as ideal [24]. We also established a direct individual consistency index (D) to assess the reliability of each questionnaire individually, using the equation:

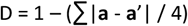

Where **a** is the five-element vector with ASRS answer scores 1, 2, 3, 5, and 6, shown for the first time, and **a’** is the vector with the scores of these answers when shown for the second time. We arbitrarily selected subjects with D ≥ 0.75 (75%) as reliable.

We studied subgroups, which are: (1) scores lower than or equal to 1, and higher than or equal to 2.5, to increase accuracy of ADHD detection; (2) ADH(+) and (−) that never were submitted to therapies; (3) inside professional niches with higher and lower ADH(+) prevalence; and (4) a randomized and stratified subsample regarding scholarship categories.

The first 206 answers were excluded for analysis because we realized online alterations on questionnaire. To obtain the dataset, contact the authors.

### Statistical analysis

To compare ASRS scores, age, and number of clinical and mental problems between ADH(+) and ADH(−) groups, we used the T-test for independent samples. For the other variables, regarded as scalar or binary, we adopted the Mann-Whitney U and Fischer’s exact tests, respectively. Correlation analyzes were performed between the ASRS score and continuous (age), scalar, and dichotomous variables using Pearson, Spearman, and point-biserial correlation methods, respectively.

Participants were clustered by professional careers. To infer if ADH(+) prevalence in each cluster is different from the expected one, we used chi-square test for categorical variables:

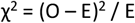

Where *O* is the total number of ADH(+) participants in the current cluster, and *E* is the number of subjects in the cluster times the rate of ADH(+)/total subjects (prevalence). For inference, we regarded one degree of freedom (if ADH(+) or not). We rank the professional niches by χ^2^ coefficient in descending order, selecting all with p < 0.1 to form two groups of professions (with higher and lower ADH(+) prevalence).

To evaluate the effect size of correlations, we used Cohen’s proposed thresholds for psychosocial sciences [25], which considers r> 0.5 as large effect, 0.3 > r ≥ 0.5 as moderate effect, 0.1> r ≥ 0.3 as weak effect, and below 0.1 as no effect. The magnitude of the difference between groups was estimated in terms of rates (0 to 100%), i.e., 1 – (smaller value / larger value), and it was considered when differences are statistically significant.

## Results

### Sample description

We collected a total of 2173 questionnaires. For ASRS questions, Cronbach’s alpha index was 0.67, regarded as suitable. Only 13 subjects left ASRS questions unanswered. The D index was 0.92 ± 0.06, with only 33 subjects excluded (D> 0.75). The total number of subjects included was 2126, with 1530 screened as ADH(−) and 596 screened as ADH(+), revealing a total prevalence of 28.03%. The mean D-index was the same between the groups (0.93 ± 0.05 for ADH(−) and 0.93 ± 0.06 for ADH(+), p = 0.273). Among genders, 70% of the answers were from females (cis or trans), corresponding to the biological sex, with an estimated participation of <1% of transgender people.

The distribution of ASRS scores (Fig 1-A) is normal (p <0.0001, Shapiro-Wilk test) with 70.96% of scores between 1.00 and 2.50, (mean 1.70 ± 0.68 std, and median of 1.66). The average age of participants was 40.30 ± 12.18 years, with normal distribution (p <0.0001, Fig 1-B). Only 4% of participants reported having technical courses as the highest level of education. Therefore, we disregarded it in subsequent analyses. Incomes and schooling distributions were non-normal (Fig 1, C and D).

**Fig 1.**
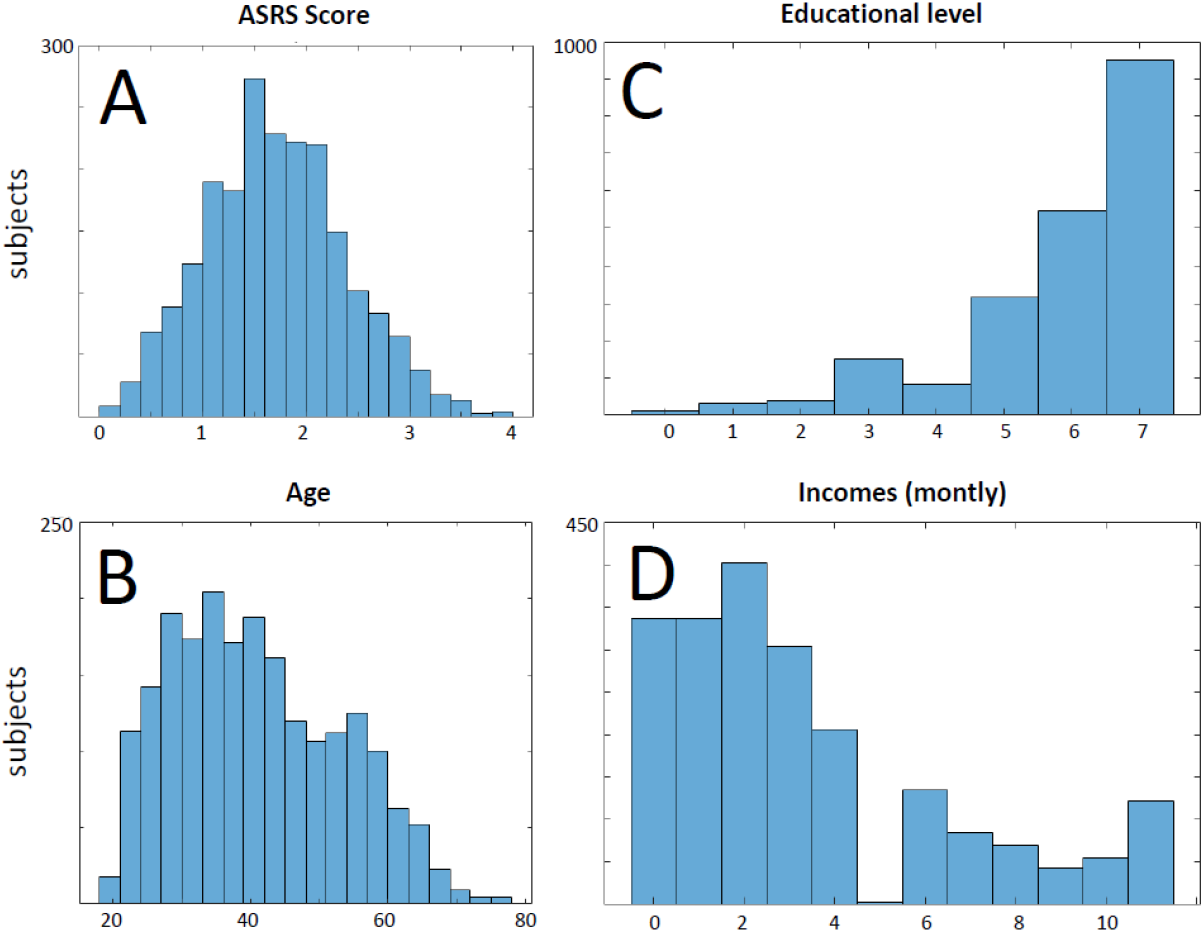
Description of the sample. The questionnaires from the 2126 selected subjects are described in terms of distributions of frequencies for (A) ASRS scores, (B) ages, (C) higher education level, and (D) financial (monthly) incomes. The classes in C are: no education (0), elementary school (1), incomplete high school (2), complete high school (3), technical degree (4), incomplete/attending graduation (5), complete graduation (6), post-graduation (7). Additionally, the classes in D are a scale of incomes with 11 levels ranging from R$ 1500.00 to “R$ 15,000.00 or more”.

About self-perception of psychological problems, 91 subjects (4.23%) stated they were ADHD (80 ADH(+) subjects by ASRS). Correcting for equalty of male and female individuals, we estimated a prevalence of 5.88%, being 7.22% and 2.95% of males and females, respectively.

### Correlation between ASRS and variables

The ASRS score in the entire sample (n = 2126) shows weak and moderate correlation with the number of self-perceived psychic problems in ADH(−) (r = 0.24, p < 0.0001) and ADH(+) (r = 0.32, p < 0.0001) groups, respectively. Age was negatively correlated with ASRS scores only in the ADH(+) group, with a weak effect (r = −0.27, p < 0.0001). All other correlations had r < 0.2, with negligible effect size.

### The effect of biological sex

The female sample is approximately 3 years older (p> 0.0001), with a statistically higher level of education by 5% (“undergraduate” class). Conversely, average female incomes are about 20% lower than those of males (p < 0.0001). Women reported more physical and psychical problems and they underwent more therapies than males. See Table 2.

**Table 2.**
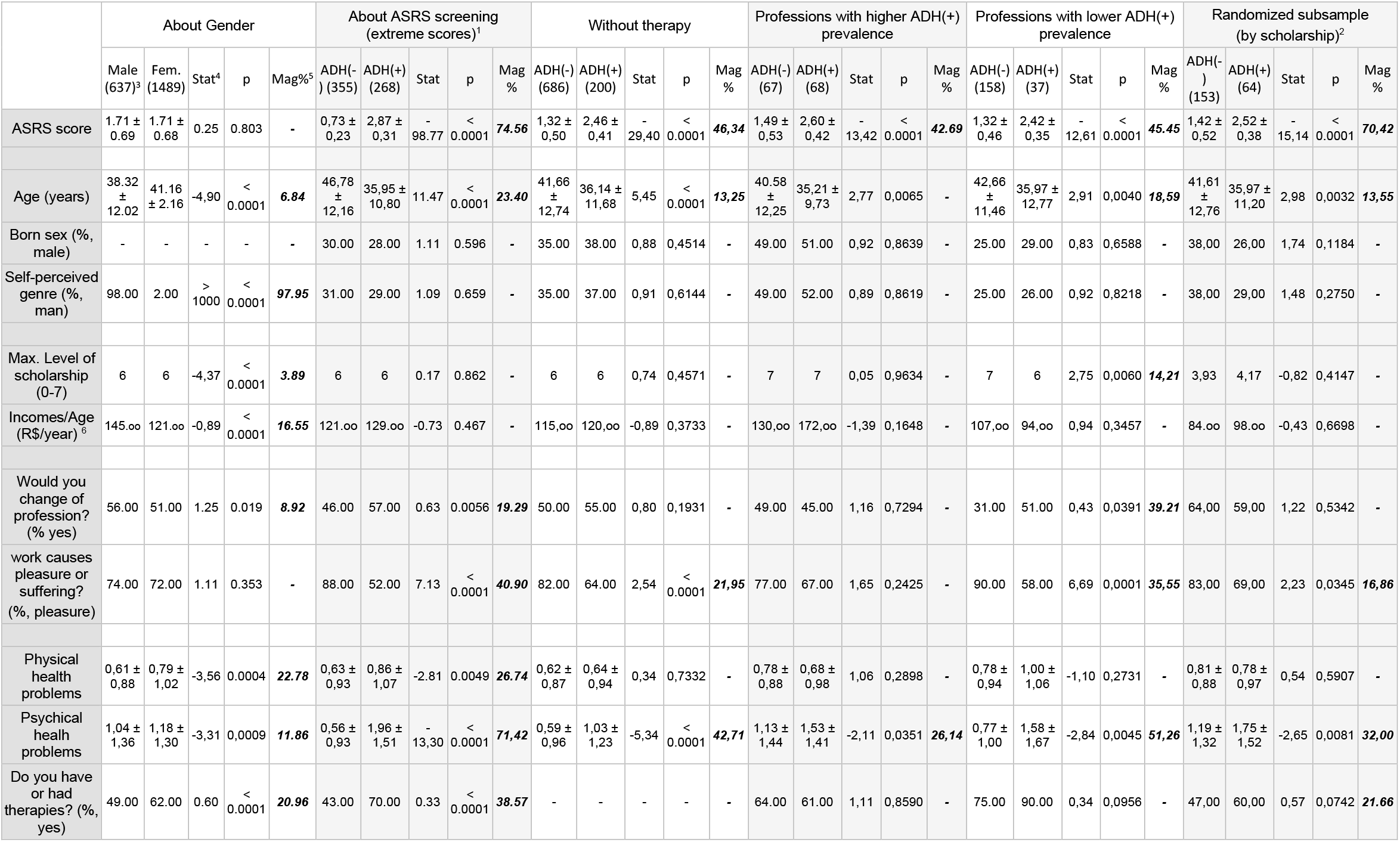
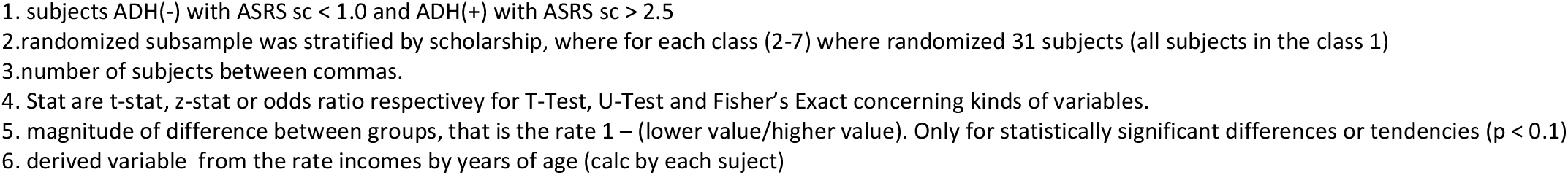
Comparisons between groups.

|  | About Gender |  |  |  |  | About ASRS screening (extreme scores)<sup>1</sup> |  |  |  |  | Without therapy |  |  |  |  | Professions with higher ADH(+) prevalence |  |  |  |  | Professions with lower ADH(+) prevalence |  |  |  |  | Randomized subsample (by scholarship)<sup>2</sup> |  |  |  |  |
| --- | --- | --- | --- | --- | --- | --- | --- | --- | --- | --- | --- | --- | --- | --- | --- | --- | --- | --- | --- | --- | --- | --- | --- | --- | --- | --- | --- | --- | --- | --- |
|  | Male (637)<sup>3</sup> | Fem. (1489) | Stat<sup>4</sup> | p | Mag<sup>5</sup> | ADH(-) (355) | ADH(+) (268) | Stat | p | Mag % | ADH(-) (686) | ADH(+) (200) | Stat | p | Mag % | ADH(-) (67) | ADH(+) (68) | Stat | p | Mag % | ADH(-) (158) | ADH(+) (37) | Stat | p | Mag % | ADH(-) (153) | ADH(+) (64) | Stat | p | Mag % |
| ASRS score | 1.71 ± 0.69 | 1.71 ± 0.68 | 0.25 | 0.803 | - | 0.73 ± 0.23 | 2.87 ± 0.31 | - 98.77 | < 0.0001 | **74.56** | 1.32 ± 0.50 | 2.46 ± 0.41 | - 29.40 | < 0.0001 | **46.34** | 1.49 ± 0.53 | 2.60 ± 0.42 | - 13.42 | < 0.0001 | **42.69** | 1.32 ± 0.46 | 2.42 ± 0.35 | - 12.61 | < 0.0001 | **45.45** | 1.42 ± 0.52 | 2.52 ± 0.38 | - 15.14 | < 0.0001 | **70.42** |
| Age (years) | 38.32 ± 12.02 | 41.16 ± 2.16 | -4.90 | < 0.0001 | **6.84** | 46.78 ± 12.16 | 35.95 ± 10.80 | 11.47 | < 0.0001 | **23.40** | 41.66 ± 12.74 | 36.14 ± 11.68 | 5.45 | < 0.0001 | **13.25** | 40.58 ± 12.25 | 35.21 ± 9.73 | 2.77 | 0.0065 | - | 42.66 ± 11.46 | 35.97 ± 12.77 | 2.91 | 0.0040 | **18.59** | 41.61 ± 12.76 | 35.97 ± 11.20 | 2.98 | 0.0032 | **13.55** |
| Born sex (% male) | - | - | - | - | - | 30.00 | 28.00 | 1.11 | 0.596 | - | 35.00 | 38.00 | 0.88 | 0.4514 | - | 49.00 | 51.00 | 0.92 | 0.8639 | - | 25.00 | 29.00 | 0.83 | 0.6588 | - | 38.00 | 26.00 | 1.74 | 0.1184 | - |
| Self-perceived genre (% man) | 98.00 | 2.00 | > 1000 | < 0.0001 | **97.95** | 31.00 | 29.00 | 1.09 | 0.659 | - | 35.00 | 37.00 | 0.91 | 0.6144 | - | 49.00 | 52.00 | 0.89 | 0.8619 | - | 25.00 | 26.00 | 0.92 | 0.8218 | - | 38.00 | 29.00 | 1.48 | 0.2750 | - |
| Max. Level of scholarship (0-7) | 6 | 6 | -4.37 | < 0.0001 | **3.89** | 6 | 6 | 0.17 | 0.862 | - | 6 | 6 | 0.74 | 0.4571 | - | 7 | 7 | 0.05 | 0.9634 | - | 7 | 6 | 2.75 | 0.0060 | **14.21** | 3.93 | 4.17 | -0.82 | 0.4147 | - |
| Incomes/Age (R\$/year)<sup>6</sup> | 145.00 | 121.00 | -0.89 | < 0.0001 | **16.55** | 121.00 | 129.00 | -0.73 | 0.467 | - | 115.00 | 120.00 | -0.89 | 0.3733 | - | 130.00 | 172.00 | -1.39 | 0.1648 | - | 107.00 | 94.00 | 0.94 | 0.3457 | - | 84.00 | 98.00 | -0.43 | 0.6698 | - |
| Would you change of profession? (% yes) | 56.00 | 51.00 | 1.25 | 0.019 | **8.92** | 46.00 | 57.00 | 0.63 | 0.0056 | **19.29** | 50.00 | 55.00 | 0.80 | 0.1931 | - | 49.00 | 45.00 | 1.16 | 0.7294 | - | 31.00 | 51.00 | 0.43 | 0.0391 | **39.21** | 64.00 | 59.00 | 1.22 | 0.5342 | - |
| work causes pleasure or suffering? (% pleasure) | 74.00 | 72.00 | 1.11 | 0.353 | - | 88.00 | 52.00 | 7.13 | < 0.0001 | **40.90** | 82.00 | 64.00 | 2.54 | < 0.0001 | **21.95** | 77.00 | 67.00 | 1.65 | 0.2425 | - | 90.00 | 58.00 | 6.69 | 0.0001 | **35.55** | 83.00 | 69.00 | 2.23 | 0.0345 | **16.86** |
| Physical health problems | 0.61 ± 0.88 | 0.79 ± 1.02 | -3.56 | 0.0004 | **22.78** | 0.63 ± 0.93 | 0.86 ± 1.07 | -2.81 | 0.0049 | **26.74** | 0.62 ± 0.87 | 0.64 ± 0.94 | 0.34 | 0.7332 | - | 0.78 ± 0.88 | 0.68 ± 0.98 | 1.06 | 0.2898 | - | 0.78 ± 0.94 | 1.00 ± 1.06 | -1.10 | 0.2731 | - | 0.81 ± 0.88 | 0.78 ± 0.97 | 0.54 | 0.5907 | - |
| Psychical health problems | 1.04 ± 1.36 | 1.18 ± 1.30 | -3.31 | 0.0009 | **11.86** | 0.56 ± 0.93 | 1.96 ± 1.51 | - 13.30 | < 0.0001 | **71.42** | 0.59 ± 0.96 | 1.03 ± 1.23 | -5.34 | < 0.0001 | **42.71** | 1.13 ± 1.44 | 1.53 ± 1.41 | -2.11 | 0.0351 | **26.14** | 0.77 ± 1.00 | 1.58 ± 1.67 | -2.84 | 0.0045 | **51.26** | 1.19 ± 1.32 | 1.75 ± 1.52 | -2.65 | 0.0081 | **32.00** |
| Do you have or had therapies? (% yes) | 49.00 | 62.00 | 0.60 | < 0.0001 | **20.96** | 43.00 | 70.00 | 0.33 | < 0.0001 | **38.57** | - | - | - | - | - | 64.00 | 61.00 | 1.11 | 0.8590 | - | 75.00 | 90.00 | 0.34 | 0.0956 | - | 47.00 | 60.00 | 0.57 | 0.0742 | **21.66** |

- 1. subjects ADH(-) with ASRS sc < 1.0 and ADH(+) with ASRS sc > 2.5 - 2.randomized subsample was stratified by scholarship, where for each class (2-7) where randomized 31 subjects (all subjects in the class 1) - 3.number of subjects between commas. - 4. Stat are t-stat, z-stat or odds ratio respectively for T-Test, U-Test and Fisher’s Exact concerning kinds of variables. - 5. magnitude of difference between groups, that is the rate 1 – (lower value/higher value). Only for statistically significant differences or tendencies (p < 0.1) - 6. derived variable from the rate incomes by years of age (calc by each subject)

### Effect of ADHD phenomenology

The proportion of male/female and men/women in the groups formed by participants with extreme ASRS scores (ADH(−) <1.0, n = 355 and ADH(+)> 2.5, n = 268) was also 70% and 30% in both ADH(+) and ADH(−) groups (table 2), respectively.

ADH(+) and ADH(−) groups are statistically the same for rate incomes/age, schooling, and number of graduations (Table 2). Regarding work suffering, 88% of the ADH(−) group said that work is a pleasure, against 52% of the ADH(+) group (p < 0.0001, see table 2). In this line, 46% and 57% people, from the ADH(−) and ADH(+) groups, respectively, would change their profession (p = 0.0056, table 2). Subjects in the ADH(+) group observedmore physical problems than those in the ADH(−) group (p = 0.0049). Mental problems are four times higher in the ADH(+) group (p < 0.0001, magnitude: 71.42%). In the ADH(+) group, 70% claimed to have already had therapy, compared to 43% of the ADH(−) group (p <0.0001).

### Effect of therapeutic intervention on ADH

There are 200 and 686 subjects in the ADH(+) and ADH(−) groups, respectively, which remain statistically equal for income/age, scholarship, and number of higher education courses. Still, these subjects did not show difference either in desire to change their professions or in the perception of physical diseases. However, psychic problems perceived by individuals in the ADH(+) group were also twice as large as the ADH(−) group (p < 0.0001), and the ADH(+) group suffered 18% more at work (p < 0.0001, table 2).

### The effect of professional niches on ADH

We found professions with both significantly higher and lower prevalence than that expected from ADH(+) subjects. The seven professions with the highest prevalence of ADH(+) (p <0.100) were: publicity (Found/Expected = 2.02, p = 0.0001), physics-degree (F/E = 3.57, p = 0.027), public defender (F/E = 2.14, p = 0.027), cinema (F/E = 2.14, p = 0.027), biology (F/E = 1.44, p = 0.056), history (F/E = 1.41, p = 0.073), medicine-mental health(F/E = 1.63, p = 0.085). See table 3.

**Table 3.**
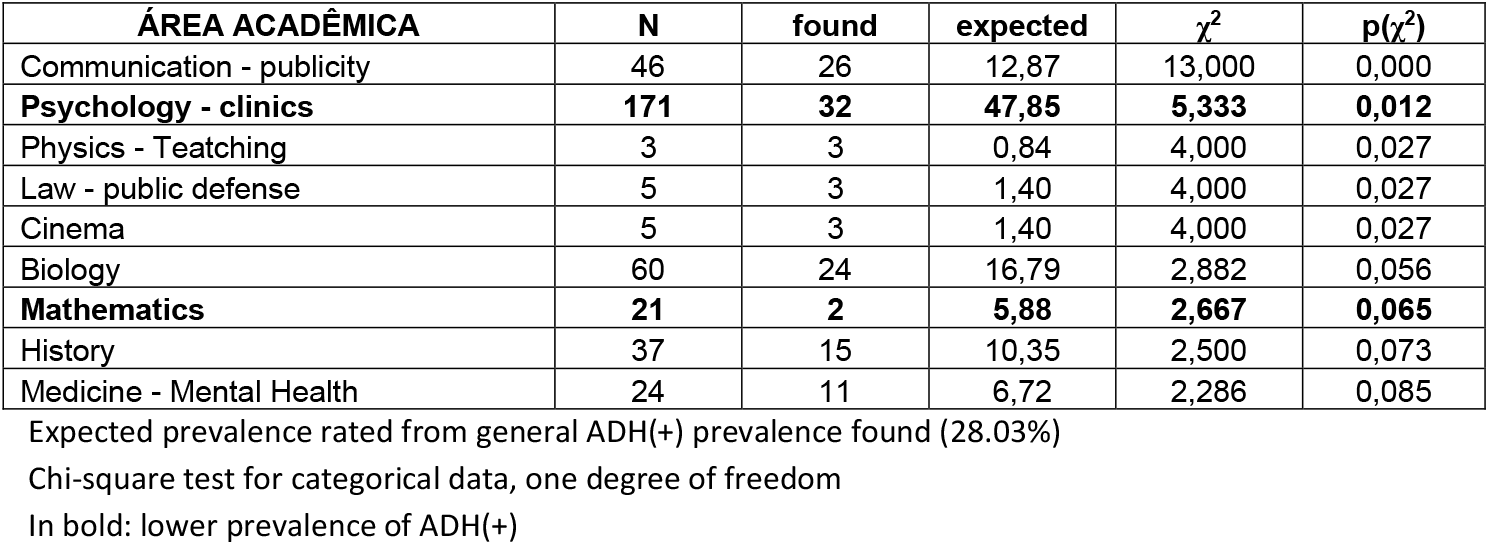
Higher and Lower ADH(+) prevalence by professional career (p <0.100)

| ÁREA ACADÊMICA | N | found | expected | $\chi^2$ | p($\chi^2$) |
| --- | --- | --- | --- | --- | --- |
| Communication - publicity | 46 | 26 | 12,87 | 13,000 | 0,000 |
| **Psychology - clinics** | **171** | **32** | **47,85** | **5,333** | **0,012** |
| Physics - Teatching | 3 | 3 | 0,84 | 4,000 | 0,027 |
| Law - public defense | 5 | 3 | 1,40 | 4,000 | 0,027 |
| Cinema | 5 | 3 | 1,40 | 4,000 | 0,027 |
| Biology | 60 | 24 | 16,79 | 2,882 | 0,056 |
| **Mathematics** | **21** | **2** | **5,88** | **2,667** | **0,065** |
| History | 37 | 15 | 10,35 | 2,500 | 0,073 |
| Medicine - Mental Health | 24 | 11 | 6,72 | 2,286 | 0,085 |

Expected prevalence rated from general ADH(+) prevalence found (28.03%) Chi-square test for categorical data, one degree of freedom In bold: lower prevalence of ADH(+)

These professions with high prevalence of ADH(+) totaled 68 ADH(+) and 67 ADH(−) subjects. Academic and economic performance also remained the same across groups (Table 2). When we evaluated subjective suffering, the groups were not different regarding occupational stress or self-perception of physical illness. However, there was also higher incidence of self-perception of psychic problems in the ADH(+) group (p = 0.0351), with a magnitude of 26.14%.

Two professions with the lowest prevalence of ADHD (p <0.100) were, in order of significance, clinical psychology (F/E = 0.66, p = 0.012) and mathematics (F/E = 0.34, p = 0.065), with a total of 31 ADH(+) and 158 ADH(−) subjects (table 3). While income/age was not different once again, ADH(+) subjects had significantly lower education than ADH(−) (p = 0.0060), equivalent to “graduation” and “post-graduation”, respectively. Regarding subjective suffering related to work stress and self-perception of physical/mental problems, ADH(+) individuals would change their profession (p = 0.0391, magnitude: 39.21%), as they related it to suffering (42% in ADH(+), versus 10% in ADH(−), p = 0.0001, magnitude: 35.55%). As in general scenarios, ADH(+) individuals also had a higher perception of psychic problems (p = 0.0045, magnitude: 51.26%). See table 2.

### The effect of ADH phenomenology on randomized stratified subsample

The rarest class was “elementary school” (31 subjects). Thus, we stratified this subsample with 217 subjects (ADH(−) = 153, ADH(+) = 64). As observed in previous comparisons, there are no differences regarding incomes and scholarship, or desire to change professions (table 2). Again, ADH(+) showed higher suffering related to work. Self-perception of clinical problems were the same between the groups, and ADH(+) mentioned more psychical problems. However, the magnitude was lower than in other scenarios (32.00%). See table 2

## Discussion

### Main characteristics of ADHD profile

The present study is a preliminary discussion about whether ADHD should be a mental disorder, according to the formal definition of DSM-5 [2], studying a general population without a priori diagnosis. If subjects with ADHD screening (+) show functionality and adaptability in specific settings, without any correlation with previous treatments, we can discuss the morbid nature of ADH as influenced by work/social stress. Despite the limitations of this study (which will be discussed later), we believe we have raised this debate with new insights. This study does not address subjects’ pathway into adulthood, although ADHD might manifest in childhood [2], but we assume that our findings are an outcome of individual history of psychosocial development.

Our core instrument to access ADHD was ASRS, a self-assessment recommended by the World Health Organization for ADHD screening. The six-question self-assessment questionnaire used here has a sensitivity of 68.7% and a specificity of 99.5% (equivalent to the 18-question version) and total classification accuracy of 97.9%, estimated regarding the DSM-IV [19]. These results show that the inventory of six questions is an ideal screening tool for research, with a very low false-positive rate. There are previous experiences using ASRS by phone or online surveys [26–29]. The reliability of answers was warranted by Cronbach’s index and ASRS re-testing amid distracting questions. The ASRS is a screening tool and we did not intend to establish ADHD diagnosis using it (so we were careful to mention ADH). Similarly, the questionnaire addresses demographic issues and people’s perceptions of themselves and their lives, without any value as (or any intention of being) a diagnostic or clinical assessment.

In our study, we analyzed the subjects in several scenarios, maximizing accuracy in ASRS screening for ADHD by selecting subjects with extreme scores, observing the effect of psychotherapy and professions with higher and lower ADH(+) prevalence. We also observed how consistent were the findings using a randomized sample stratified by schooling, which did not have an original distribution of frequency that was representative of the overall population.

In nearly all scenarios (except for professions with lower ADH(+) prevalence), functionality in terms of academic and economic performance was statistically the same between people with (+) and (−) screening for ADHD. According to the ASRS screening in the universe studied, these findings are counterintuitive in light of our premises regarding ADHD. However, they clearly indicate that ADHD symptoms are not related with what has historically been established [17, 18]. It does not seem to be a bias generated by the schooling profile of the sample, as the randomized/stratified scenario confirms the results. One explanation for our results might be that outcome studies that built a morbid profile for ADHD have been conducted with samples of institutionalized people with well-established mental disorders or with history of school problems, and not with the overall population through a blind study, with participants who are naïve regarding the objectives of the study. We have not identified previous studies with designs compatible with ours.

On the other hand, in almost all scenarios, the perception of psychic (and even physical) problems as well as the report of labor dissatisfaction, which would denote the ability of people to adapt to life stressors, was worse in ADH(+) people. However, in the setting of professions with higher ADH(+) prevalence, this phenomenon was substantially mitigated: there is no longer evidence of labor stress, of higher perception of clinical illness, nor of higher frequency of therapies undergone. Additionally, the magnitude of differences between groups regarding perception of psychic problems is the lowest among all scenarios. In this particular case, if we correct the p-value using the Bonferroni method, the difference is no longer significant (multiplying by six, the number of studied scenarios). On the other hand, in professional niches with lower ADH(+) prevalence, the ADH(+) group shows once again a pattern of higher suffering, and indirectly, lower schooling compared to the ADH(−) group. These results corroborate the idea that the disability clearly observed in the ADH(+) group would primarily result from the influence of socio-cultural stressors, in this case related to labor and its context of social relationships, which are a cardinal dimension of human life.

Publicity was the professional niche with the highest prevalence of ADH(+) people, with absolute significance in probability of difference between groups (p = 0.0001, or p = 0.0096, corrected by the Bonferroni method). It is obvious that cognitive work profile as well as organization of the environment and work relations in Publicity is quite different from teaching mathematics or exercising clinical psychology. ADH(+) people probably identify a higher compatibility between their personality and skill profiles and certain professional profiles, in which the morbid effects currently related with ADHD are significantly mitigated.

As a parameter for a comparative discussion, we studied the differences between male and female genders in the same dimensions. With the historical knowledge that there are differences between genders regarding income in the Brazilian setting [30] and incidence of psychic suffering secondary to stress [31] although gender is not considerable as a morbid factor nor a disorder in itself, a qualitatively similar profile of differences was found between male and female genders and between ADH(+) and ADH(−) scenarios. Women manifested lower dissatisfaction with their profession; however, they showed higher perception of physical and psychic problems, and higher frequency in undergoing therapies. In accordance with national statistics, women showed a significantly lower economic performance, even having higher educational level (this finding is an indicator of quality in our data). Similar to being a woman, having ADHD profile might be understood as a risk factor for illness due to social and cultural stress and due to stigmatization.

The inverse relationship between age and ASRS scores was one of the most typical results, in any of the five studied scenarios. This relationship is more evident in the ADH(−) group, and is not significant in the ADH(+) group, thus showing dimensionality between typical people. This result suggests that ADHD symptomatology might be related to a slower development of cognitive functions [10,34,35].

Finally, another issue observed was the great prevalence of ADH(+) people regarding current epidemiological studies [32, 33], which was 28.03%. The first thing that comes to mind is related to ASRS quality, as well as the method of application. However, a possible explanation for this is related to the nature of the recruitment of participants in this study, which shall be discussed below.

### Methodological considerations and limitations

One evident limitation of this study was the non-randomization of subjects during recruitment. An on-demand sample is subject to several biases, which are identified through distributions of frequencies not compatible with the original population. As we recruited 2126 subjects, we were able to perform a subsampling with randomization and stratification by schooling level, which mitigated the sampling bias in the inference of results about functionality and adaptability in the overall population.

The 28% prevalence of ADHD found here might have resulted from the on-demand sampling, related to the public title of the survey (“Relationship between Behavior, Health, and Working Life”), which is more striking for people who experience problems or suffering at work and in overall health. However, since we have a strict report of ASRS scores, as this is a validated instrument, and our inference derived from a randomized/stratified subsample, we can assume that there was no impact on the quality of the results. Overall, the ADHD prevalence observed in several studies has varied greatly [36–39], reaching 16.4% in a survey over the phone using DSM-IV criteria [26].

A pertinent question in this study is the appropriateness of p-value correction as we deal with scenarios of multiple comparisons. In preliminary studies of exploratory nature, which must be especially sensitive to any potential differences, type-II errors might be more harmful that type-I errors, which leads to our questioning of p-value correction [40, 41]. Moreover, the most conservative hypothesis in this study, particularly, is H1 rather than H0 (there is no similarity between groups). Therefore, it is also more conservative to admit type-I rather than type-II errors. Regarding the selection of professional niches by ADH(+) prevalence, χ^2^ statistics and p-values were not particularly considered for direct inference.

### Conclusion and Final considerations

While the vast majority of mental disorders have been considered for centuries, the first systematization of the ADHD phenomenon as a clinical disorder was only performed in 1902, considering the “defect of moral control” as a manifestation of “some morbid physical condition”[42]. Even though ADHD is a global perception [43], its estimated prevalence fluctuates too much between different regions in the world in studies that use the same diagnostic guidelines [32], which should be seen as evidence of a nosological inconsistency. In fact, ADHD is a theoretical model for a presumed disorder.

Indeed, ADHD is a well-defined entity biologically. It was related to slower maturation of the prefrontal cortex with impaired selective and sustained attention and behavioral inhibition [10,34] associated to the right hemisphere prevalence [9, 44], which could explain a higher expressiveness of creative and imagery skills (necessary for Publicity), along with verbal and analytical disabilities (cardinal for psychology and mathematics) [45].

However, the fact that it is a biologically entity is far from having a presumed psychopathological nature. Our findings in this preliminary study consistently point to an alternative interpretation of attention deficit and hyperactivity phenomenology not as a disorder itself but primordially as a different cognitive style under circumstantial stress, manifesting suffering because of that stress. Thus, these particularities of the cognitive functioning of a minority of people with ADH profile would become secondarily maladaptive when confronted with the operative and moral norms of society determined by the predominant cognitive style in the statistical majority of human population. In the future, ADH might be viewed only as an element of human diversity whose subjects today suffer from social stress (e.g. women) because they are a true minority. We hope that further studies will better answer the hypotheses raised here.

## Data Availability

All data produced are available as supplementary material

## Acknowledgements

The authors are grateful to Oswaldo Cruz Foundation and Faculdade Arthur Sá Earp Neto for supporting this research, and also to all people who participated in the project. They also thank to Mrs C. Fernandes for implementing the questionnaire in Google Forms. On behalf of all authors, the corresponding author states that there is no conflict of interest.

## Notes

### Competing Interest Statement

The authors have declared no competing interest.

### Funding Statement

This study did not receive any funding

### Author Declarations

This work and the procedures adopted were duly approved by the Research Ethics Committee of Fernandes Figueira National Institute of the Woman, Child and Adolescent Health (CEP-IFF/FIOCRUZ), registered by CAAE: 80765917.7.0000.5269, under national laws and in accordance with the ethical standards laid down in the 1964 Declaration of Helsinki and its later amendments.

